# Estimates of the eligible population for Australia’s targeted National Lung Cancer Screening Program, 2025-2030

**DOI:** 10.1101/2024.07.02.24309739

**Authors:** Stephen Wade, Preston Ngo, Yue He, Michael Caruana, Julia Steinberg, Qingwei Luo, Michael David, Annette McWilliams, Kwun M Fong, Karen Canfell, Marianne Weber

**Affiliations:** The Daffodil Centre, The University of Sydney, a joint venture with Cancer Council NSW, Woolloomooloo, NSW, Australia; School of Medicine and Dentistry, Griffith University, Gold Coast, QLD, Australia; Fiona Stanley Hospital, Perth, WA, Australia; Prince Charles Hospital, Chermside, QLD, Australia; The University of Queensland Thoracic Research Centre/The Prince Charles Hospital, Brisbane, QLD, Australia

## Abstract

Australia’s National Lung Cancer Screening Program will commence in July 2025, targeted at individuals aged 50-70 years with a 30 pack-year smoking history, who either currently smoke or have quit within the last 10 years. We forecasted the number of screening-eligible individuals over the first 5 years of the program using data from the 2019 National Drug Strategy Household Survey, and the 2022 Australian Bureau of Statistics population projections. Multiple imputation integrated with predictive modelling of future or unmeasured smoking characteristics was used to address missing data and simultaneously, to project individuals’ smoking histories to 2030. In 2025, 930,500 (95% prediction interval 852,200-1,019,000) individuals were estimated to be eligible, with the number meeting the criteria declining slightly over the years 2025-2030 in all Australian jurisdictions. Overall, 26-30% of those eligible will have quit smoking, and 70-74% will currently smoke. These estimates can be used in resource planning and as an indicative denominator to track participation rates for the program over time.

## Introduction

Australia is preparing to launch a targeted National Lung Cancer Screening Program starting July 2025.^1^ The program will be open to individuals aged 50-70 years with a 30 pack-year smoking history (equivalent to a pack of 20 cigarettes a day for 30 years), who either currently smoke or have quit within the last 10 years. Participants will be referred from primary care to receive a low dose computed tomography scan of the chest every two years. Estimates of the number of people who qualify for lung cancer screening are important for understanding the impact of screening on resource utilisation across the screening and assessment pathway, and for program evaluation over time. Since there is no comprehensive collection of data on smoking behaviours at a population level, we forecasted the number of screening-eligible individuals over the first 5 years of the program using a statistical modelling approach.

## Methods

The number of individuals who meet the lung cancer screening age and smoking history eligibility criteria over the period 2025-2030 was estimated using data from the 2019 National Drug Strategy Household Survey,^2^ and the 2022 Australian Bureau of Statistics population projections.^3^ Multiple imputation integrated with predictive modelling of future or unmeasured smoking characteristics was used to address missing data and simultaneously, to project individuals’ smoking histories to 2030.

For each respondent, we required calculations of, 1) The age that they started smoking daily and the age that they stopped (if they stopped); 2) The duration that they had smoked or will smoke (if they start); 3) The lifetime average number of cigarettes per day, both factory made and roll-your-own (past and future). Pack-years were calculated as the number of years smoked multiplied by the number of cigarette packs smoked per day, assuming 20 cigarettes per pack.

Values of key smoking-related variables were imputed/predicted using demographic factors and other covariates (see Supplementary Material). Age at smoking initiation and smoking duration were modelled with accelerated failure time models and all remaining variables were modelled with random forests. For each state/territory and year, we predicted the proportion of the population eligible for screening using logistic regression. Weights were adjusted to sum to the effective sample size proposed by Kish^4^ and 95% prediction intervals were estimated by sampling regression parameters from the asymptotic distribution in each imputed dataset, then pooling all predictions of the eligible population.

Detailed methods are reported in the Supplementary Material.

## Results

Overall, between 12.8-14.1% of the Australian population aged 50-70 years were estimated to meet the National Lung Cancer Screening Program age and smoking criteria in the first 5 years of the program (30-33% of those with a history of smoking). In the first year, 930,500 (95% prediction interval 852,200-1,019,000) individuals were estimated to be eligible, with the number meeting the criteria in each state and territory declining slightly over the years 2025-2030 (see Table). Overall, 26-30% of those eligible will have quit smoking, and 70-74% will currently smoke.

**Table.** Estimated number (95% prediction interval) of individuals who meet the age and smoking history eligibility criteria for the National Lung Cancer Screening Program in Australia, 2025-2030.

| **State** | **2025** | **2026** | **2027** | **2028** | **2029** | **2030** |
| --- | --- | --- | --- | --- | --- | --- |
| ***New South Wales*** | 251,400 (215,600-291,600) | 249,800 (215,200-288,300) | 252,500 (217,800-291,100) | 251,800 (215,900-291,200) | 240,400 (206,100-277,600) | 235,700 (203,600-272,400) |
| ***Victoria*** | 203,000 (170,200-240,900) | 198,000 (167,200-231,100) | 197,000 (166,100-228,100) | 197,400 (164,600-232,000) | 189,600 (160,300-221,000) | 187,100 (157,600-219,400) |
| ***Queensland*** | 233,700 (198,100-271,300) | 235,500 (200,200-272,100) | 235,000 (200,000-273,800) | 232,200 (198,400-270,400) | 232,000 (197,700-269,000) | 228,700 (195,500-266,800) |
| ***Western Australia*** | 111,300 (86,100-139,500) | 111,500 (85,700-138,700) | 116,500 (94,200-143,100) | 114,000 (91,000-141,000) | 114,000 (90,400-142,200) | 109,100 (85,600-137,500) |
| ***South Australia*** | 84,900 (71,200-99,200) | 82,600 (68,700-97,500) | 81,400 (68,100-95,700) | 78,800 (65,200-93,900) | 76,600 (63,600-90,400) | 76,100 (62,400-91,700) |
| ***Northern Territory*** | 10,200 (7,700-13,100) | 10,200 (7,500-13,200) | 10,100 (7,400-13,200) | 9,800 (7,200-12,900) | 9,500 (7,000-12,600) | 9,100 (6,500-12,000) |
| ***Tasmania*** | 26,600 (19,700-34,400) | 27,400 (20,700-35,500) | 26,700 (19,400-35,200) | 26,700 (19,700-35,000) | 25,800 (18,800-34,000) | 25,700 (18,800-33,700) |
| ***Australian Capital Territory*** | 9,400 (5,600-14,200) | 9,600 (5,800-14,400) | 9,500 (5,400-14,500) | 8,600 (5,200-13,100) | 8,100 (4,800-12,100) | 7,600 (4,300-11,800) |
| ***Total*** | 930,500 (852,200-1,019,000) | 924,400 (845,000-1,012,400) | 928,800 (852,300-1,006,100) | 919,200 (844,000-996,900) | 896,000 (831,100-966,400) | 879,100 (811,500-946,600) |

## Discussion

Using individual-level, nationally representative data on smoking status, duration, intensity and years since quit smoking (if quit), we projected smoking characteristics for the Australian population up to the year 2030 by age. By applying the age and smoking eligibility criteria for the National Lung Cancer Screening Program, these estimates represent the total number of individuals who may qualify for screening in any given year.

We find that around a third of the ever-smoking population would be eligible for lung cancer screening, which is in the ballpark of earlier, more conservative estimates based on varied screening eligibility criteria using data from cohort studies (estimated 17.9%-28.5%).^5,6^

The eligibility criteria for Australia’s screening program preferentially selects individuals who currently smoke. We report that over 7 in 10 people who will be eligible for lung screening would also benefit from smoking cessation support.

These estimates are subject to several caveats and limitations. They do not account for other screening exclusions, such as symptoms suggestive of lung cancer, nor smoking-related mortality, so the true number of people eligible is likely to be somewhat lower than the estimates presented here. Moreover, the prediction intervals account for uncertainties relating to missing data and model parameters, but not for uncertainty in the choice of modelling approach. Future work can extend the methods to address these limitations.

In order to evaluate state-and territory-based resource requirements for the program, these estimates can be used to model a range of key performance indicators observed in the screening programs of other jurisdictions (e.g., Canada,^7^ UK,^8^ USA^9^) such as anticipated screening participation and adherence rates, as well as the resource implications for nodule management, actionable incidental findings, smoking cessation support, and lung cancer diagnoses. These estimates can also be used as an indicative denominator to track participation rates for the program over time.

## Data Availability

All data produced in the present work are contained in the manuscript using the National Drug strategy Household Survey available from the AIHW: https://www.aihw.gov.au/about-our-data/accessing-data-through-the-aihw/data-on-request

## Acknowledgements

The authors acknowledge the Australian Institute of Health and Welfare who supplied data from the National Drug Strategy Household Survey to be used in this analysis.

## Supplementary Material to

### Data Sources

The National Drug Strategy Household Survey 2019 was used to ascertain smoking and explanatory variables. ^1^

#### Smoking variables

Observed smoking characteristics for each respondent were ascertained from the following questionnaire items:

- D5: Smoked 100 cigarettes in a lifetime (Y/N).
- D6: Smoke daily now, in the past, or never.
- D7: Age stopped smoking daily (YY).
- D8: Age started smoking daily (YY).
- D12: Number of factory-made cigarettes smoked currently per day.
- D13: Number of roll-your-own smoked currently per day.

Respondents were categorise as having ‘never smoked’ (daily), having ‘formerly smoked’, or ‘currently smoke’ based on D5 and D6.

#### Explanatory variables

The key explanatory variables in the analysis were birth year, sex, and state of residence. For multiple imputation, we considered the following short list of predictor variables: country of birth, socio-economic indexes for areas (SEIFA) ‘Index of Relative Socio-Economic Advantage and Disadvantage’; household income; main language spoken at home; employment status; type of household; alcohol summary status, and; self-reported previous diagnosis or treatment for each of; heart disease, hypertension, asthma, depression, anxiety, and cancer.

The 2022 Australian Bureau of Statistics population projections (medium series) for people aged 50 to 70 years, inclusive, was used as the denominator for calculations of the number of people eligible for screening in the years 2025-2030.^2^

### Multiple imputation and predictive modelling of future or unmeasured smoking characteristics

For each respondent, we required calculations of:

1. The age that they started smoking daily.
2. The duration that they had smoked or will smoke (if they start) assuming they do not die first.
3. The lifetime average number of cigarettes per day, both factory made and roll-your-own, they had smoked, or will smoke (if they start).

We censored the events of starting and stopping smoking 2 years prior to the survey. We made this choice to account for either experimentation which is subsequently recanted (e.g. short periods of daily smoking at early ages that would likely be reported as never smoking in later years) or relapse (short periods of cessation of smoking followed by resumption of daily smoking). We have the following cases of censoring by smoking status:

- *Never smoked or started smoking within 2 years prior to survey*: The age at smoking initiation was left-censored at the age surveyed minus 2 years. The duration of smoking was missing. The cigarettes per day reported at the time of the survey was zero.
- *Formerly smoked and quit at least 2 years prior to the survey*: The cigarettes per day reported at the time of the survey was zero, and either:
  - the age-started and the duration of smoking were both observed; or;
  - the age-started was not recorded therefore duration was left-censored by age-stopped.
- *Currently smoke for at least 2 years and/or quit less than 2 years prior to survey*: The duration was right-censored at the age-surveyed minus age-started minus 2 years, and either;
  - the respondent did not report having quit and their cigarettes per day was either recorded or missing, or;
  - the respondent did report having quit, and their cigarettes per day was missing.

Multiple imputation was performed using Multiple Imputation by Chained Equations (MICE).^3,4^ We chose to generate 8 imputed values for each missing value or censored time-to-event (age-started and duration). Each chain was run for 30 iterations. According to S7.1 of White, et al. (2011), using 8 chains and allowing for a 5% loss of efficiency, a fraction of missing information of up to 0.4 is acceptable. The largest fraction for P(eligible) in our analysis was 0.396.^5^MICE requires that each key variable with missing values has a ‘fully conditional specification’; i.e. that we specify the relationship between explanatory covariates (predictors) and the (missing) outcome. We constructed a short list of variables in the data set that might be related to our analysis variables (age-started, age-stopped, etc). The short list contained:

- country of birth (Australia, New Zealand-Oceania, United Kingdom, Europe, South-East Asia, Other Asia, North Africa and Middle East, Sub-Saharan Africa, Americas, Other) - N.B. ‘Other’ was treated as missing due to small numbers leading to failures of convergence within flexsurv;^4^
- Socio-Economic Indexes for Areas (SEIFA) ‘Index of Relative Socio-Economic Advantage and Disadvantage’;
- household income;
- main language spoken at home (English, language other than English);
- employment status (currently employed, student, unemployed/looking for work, solely engaged in home duties, retired or on a pension, volunteer/charity work, other);
- type of household (single with dependents, couple with dependents, parents with non dependent children, singles without kids, couple without kids, other);
- alcohol summary status (daily, weekly, less than weekly, ex-drinker [not in last 12 months], never drinker [full glass]), and;
- self-reported previous diagnosis or treatment for heart disease; hypertension; asthma; depression; anxiety, and; cancer (Y/N ‘have been diagnosed and/or treated’).

To select predictors, the *quickpred* function from the MICE package was used with the parameter related to the minimum proportion of usable cases equal to 0.3, and the parameter for the minimum correlation equal to 0.1.

Age-started and duration were modelled using parametric ‘accelerated failure time’ models with a cure fraction for age-started (i.e. some people will never smoke). These models were embedded into the multiple imputation loop (see below). The underlying distributions were Burr Type XII and Weibull for age-started and duration, respectively. The cure fraction for age-started was a function of state and sex and an age-surveyed spline with two-degrees of freedom; while the ‘accelerated failure time’ variables were selected as above (with the exception for duration where an age-surveyed spline with two-degrees of freedom was added). Censoring for observed values was incorporated using the rules described above for each category of respondent (‘never smoked’, etc.).

The mean and entropy of the age-started and duration per iteration and chain were plotted to assess convergence.

#### Burr Type XII cure distribution

The 3-parameter Burr Type XII distribution has scale α and shape parameters *c* and *k*:

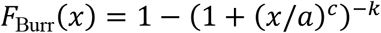

see Equation (3) in Tadikamalla (1980).^6^ To construct a cure model, a cure fraction *C* is introduced and thus:

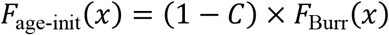

#### Kish’s effective sample size

The expression for Kish’s effective sample size given weights *C*is:

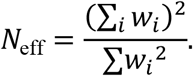

#### Accelerated failure time model within MICE

The following steps summarise the fitting and imputing for an accelerated failure time model within the MICE loop:

1. Train the model on the observed and censored events (via flexsurv::flexsurvreg)
2. Draw a value of the parameters using asymptotic distribution of the MLE (obtained via the Hessian of the likelihood).
3. Get lower and upper bound on CDF for each missing or censored observation via flexsurv::summary.flexsurvreg(type=“survival”) using the parameters drawn in 2.
4. Draw uniformly between the lower and upper bound on the CDF for each censored event or missing value.
5. Get the predicted time for each censored or missing data via inverse CDF using quantiles from 4 (and parameter values from 2).

### Confidentialised age

Ages greater than 80 years are confidentialised in the NDSHS. A penalised composite link model was used to estimate the distribution of age in single years,^7^ using the ungroup R package,^8^ for each imputed data set.

### Estimating eligibility

#### Lifetime average number of cigarettes

To estimate *lifetime* smoking history and hence eligibility, which was not fully captured by the survey, we predicted the ‘lifetime average’ number of cigarettes per day for all respondents given complete (imputed) datasets. Similar to the smoking history model developed by CISNET for the U.S. population,^16^ we assumed that smoking intensity (cigarettes per day) may change over time in the years following the age of smoking initiation, to stabilise at age 30 years. We also account for a potential reduction in smoking intensity at ages older than 60 years.

Thus, it was assumed that ‘lifetime average’ number of cigarettes was identical to the number of cigarettes per day recorded in the survey, *if*: the respondent smoked at the time of the survey and were either;

- aged 30 to 60 years, or;
- were aged less than 30 years and fewer than 10 years younger than the mid-point of their age-started and age-stopped smoking.

For respondents who did not satisfy these criteria or for whom the ‘lifetime average’ cigarettes per day was not observed, a random forest was used to predict the ‘lifetime average’. Specifically, the ‘lifetime average’ for a person P was assumed to have the same distribution as for people with

- observed ‘lifetime average’ cigarettes-per-day,
- whose age at the survey was equal to the mid-point of P’s age-start and age-stopped (or the median age [capped at 60 years] of their smoking period),

conditional on any other explanatory variables.

For example, for a respondent aged 65 years at the time of the survey who reported current smoking from the age of 20, smoking duration was imputed/predicted to determine potential age stopped (e.g. at age 70). Lifetime average cigarettes per day was then predicted based on the mid-point of their smoking period (i.e. that of a respondent aged 45 years). Whereas, if imputed/predicted age-stopped was age 90, then the mid-point of their smoking period would be that of a respondent aged 55 years old.

#### Pack-years and years-since-quit

To determine the pack-years and years-since-quit in future years for each respondent in each imputed data set, the predicted value for any censored event was used along with their lifetime average cigarettes per day. For example, a hypothetical respondent with a 30 pack-year history, who smoked 1 pack per day, would continue accruing pack-years in each year after the survey until the predicted age-stopped (see above); then they would accrue years-since-quit.

#### Post-stratification weights and probability of eligibility

To align the National Drug Strategy Household Survey sample with the general population, post-stratification weights for the survey were estimated for each calendar year using the ABS projected age, sex, and state of residence, for each imputed data set.

Finally, we modelled the probability of eligibility in each imputed data set, using logistic regression with the glm() function in R. The predictors were all combinations of state and year. Weights used in glm() were adjusted to sum to Kish’s effective sample size for each year (see above). This ensures better coverage of the prediction intervals compared to weights that sum to the number of samples.

### Pooled estimate

To obtain combined estimates from all imputed data sets; we drew values from the asymptotic distribution of the parameters of each logistic regression model (given the imputed data). Each of these values were then used to draw a prediction for the number of eligible in each state and year. We then used the 2.5th and 97.5th percentiles of the sample obtained (for each state and year) to describe the 95% prediction interval, along with the mean of the sample as a point estimate.

### Model Performance and Results

The proportion of variables missing or censored that were included in any model and had at least one missing value is presented in the Supporting Table. Model performance and results are presented in Figures S1-S3.

**Supporting Table:** Percent of variables missing or censored that were included in any model and had at least one missing value.

|  | Percent missing | Percent censored |
| --- | --- | --- |
| Had 100 cigarettes (D5) | 0.4% |  |
| Daily smoking (D6) | 1.2% |  |
| Age started (D8) | 1.2% | 60.6% |
| Duration (D7-D8) | 1.5% | 73.7% |
| Factory-made cigarettes per day | 6.7% |  |
| Roll-your-own cigarettes per day | 10.9% |  |
| Country of birth | 7.9% |  |
| Household income | 29.6% |  |
| Main language spoken | 2.8% |  |
| Employment status | 4.0% |  |
| Type of household | 1.1% |  |
| Alcohol summary status | 0.9% |  |
| Self-reported previous diagnosis or treatment: Heart disease | 6.2% |  |
| Self-reported previous diagnosis or treatment: Hypertension | 4.5% |  |
| Self-reported previous diagnosis or treatment: Depression | 5.7% |  |
| Self-reported previous diagnosis or treatment: Anxiety | 6.1% |  |
| Self-reported previous diagnosis or treatment: Cancer | 16.5% |  |

\*NB SEIFA was 100% observed

**Figure S1a:**
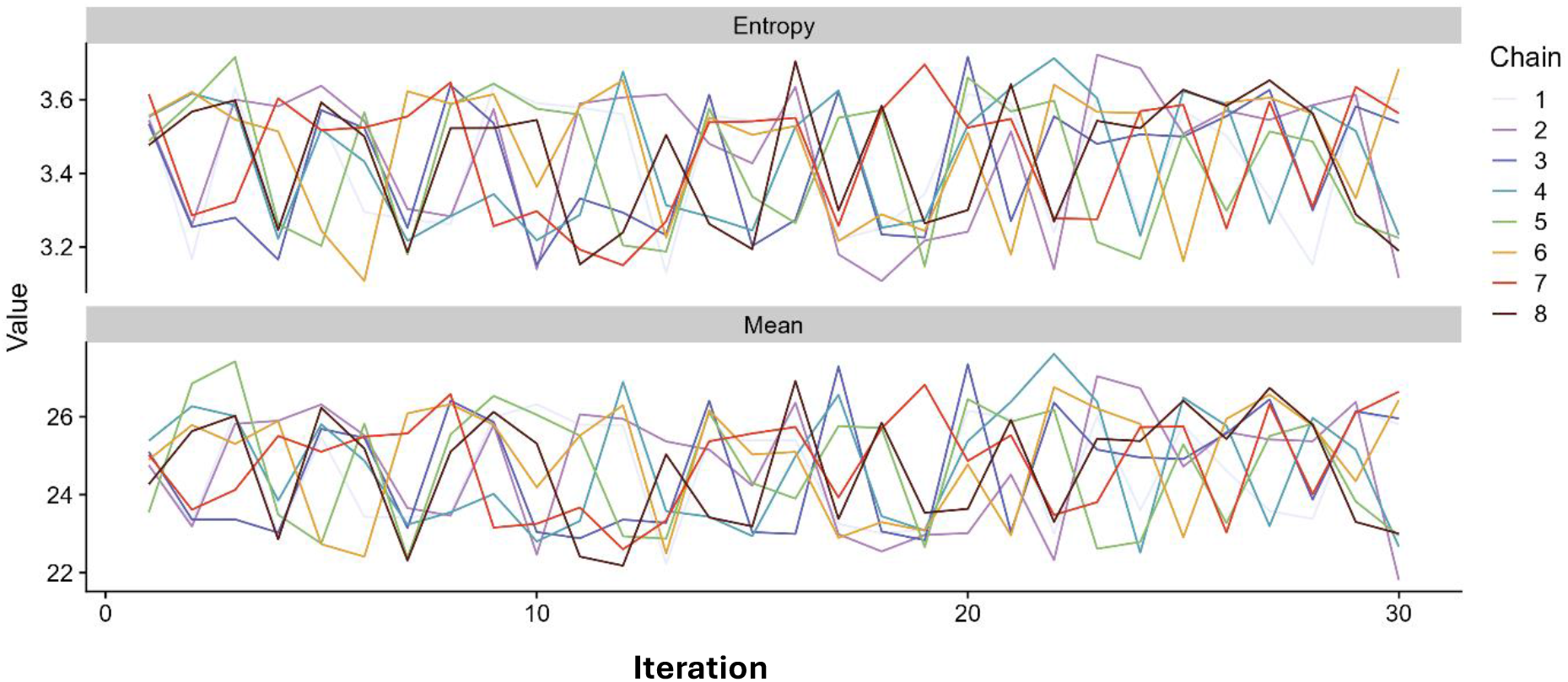
Traceplot of summary statistics for imputed age-at-started

**Figure S1b:**
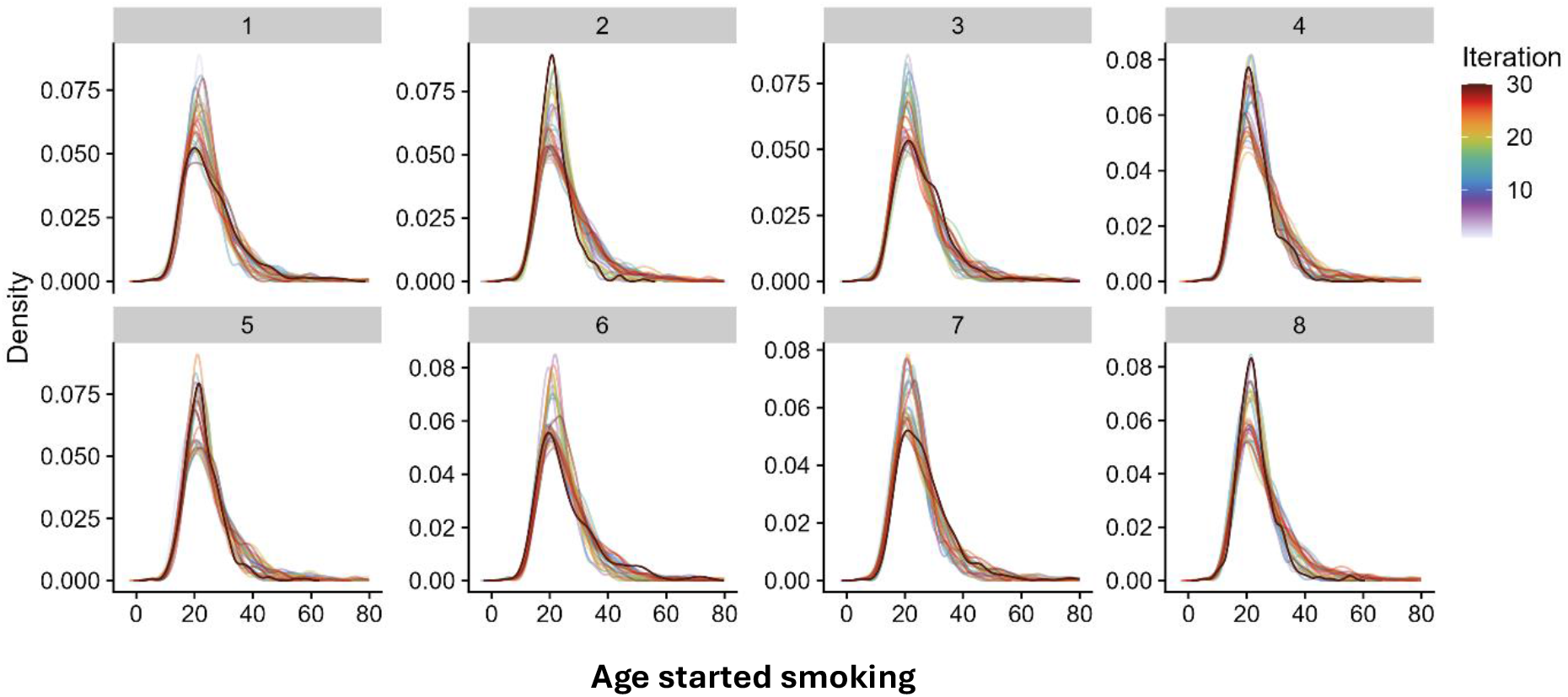
Smoking density of imputed age-at-started smoking for each chain

**Figure S2a:**
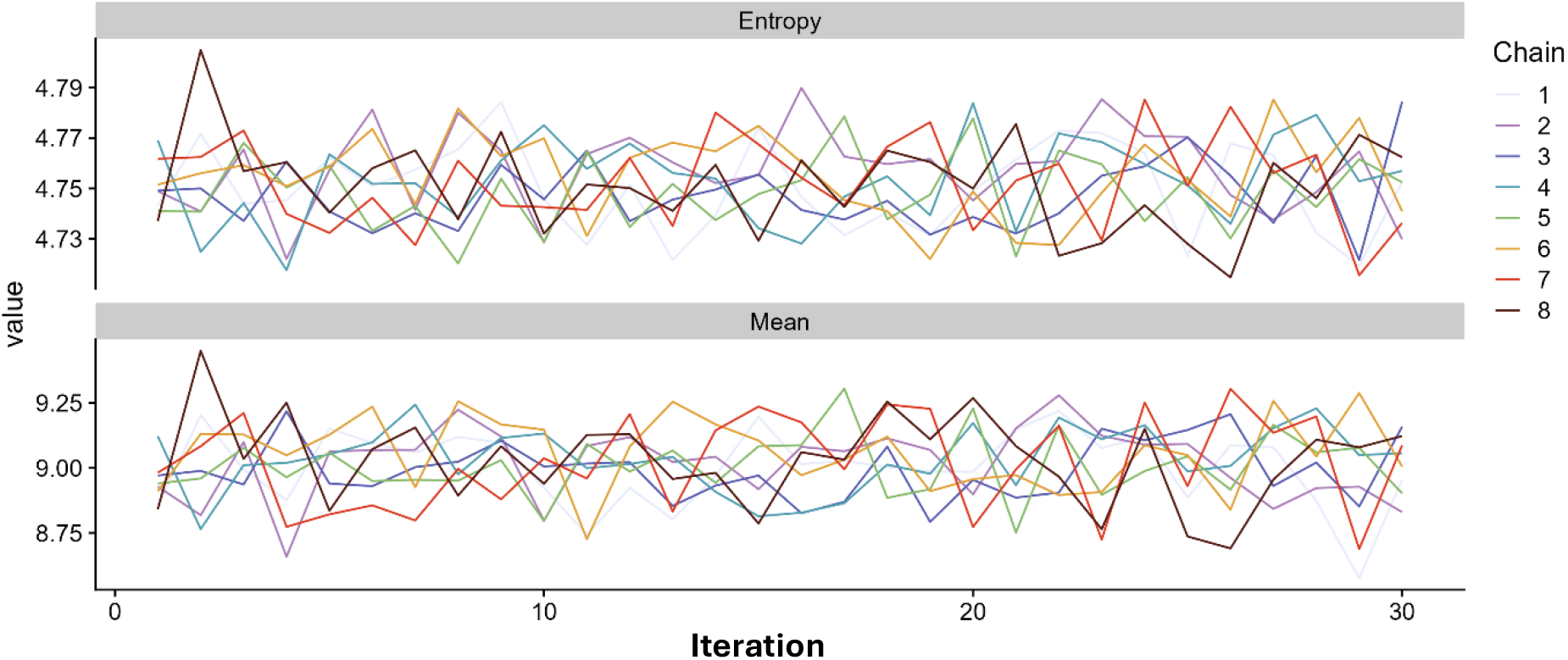
Traceplot of summary statistics for imputed duration of smoking

**Figure S2b:**
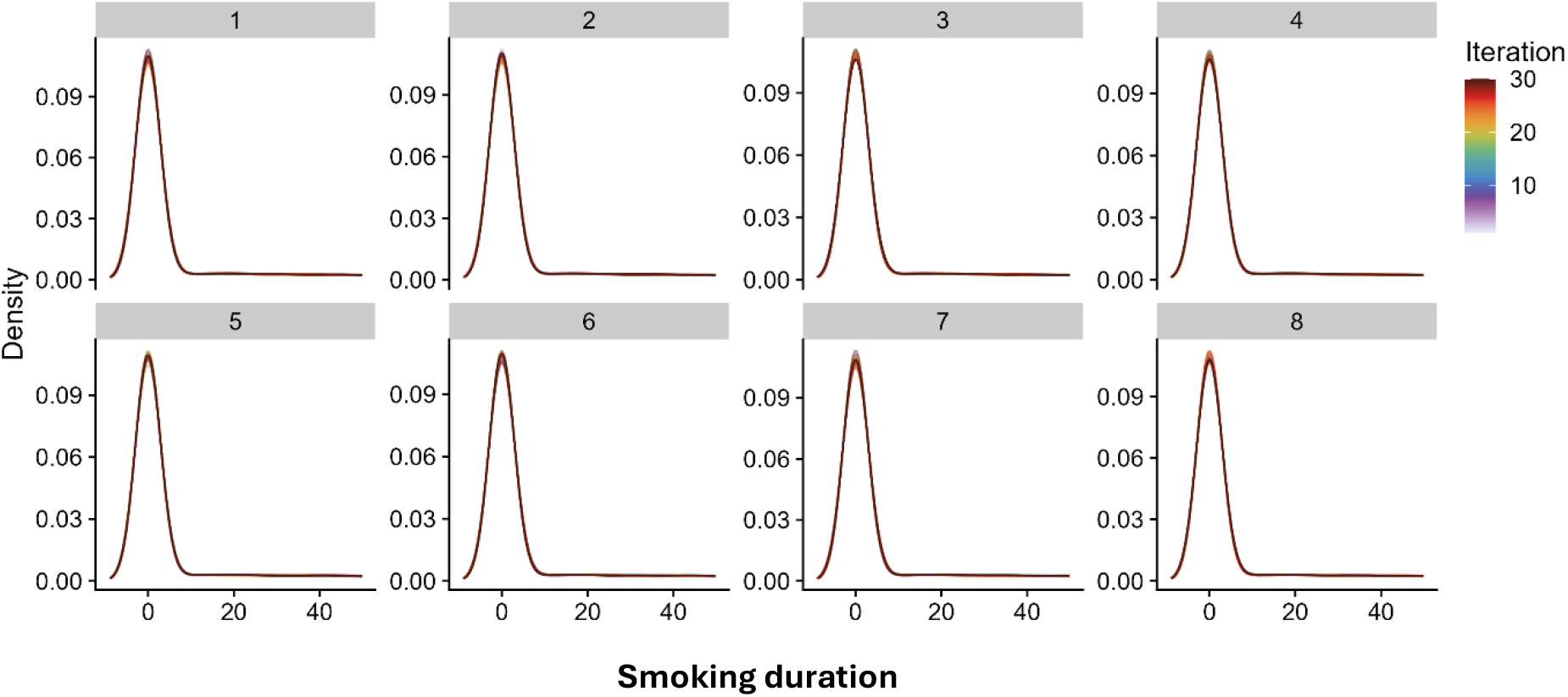
Density of imputed duration of smoking for each chain.

**Fig S3:**
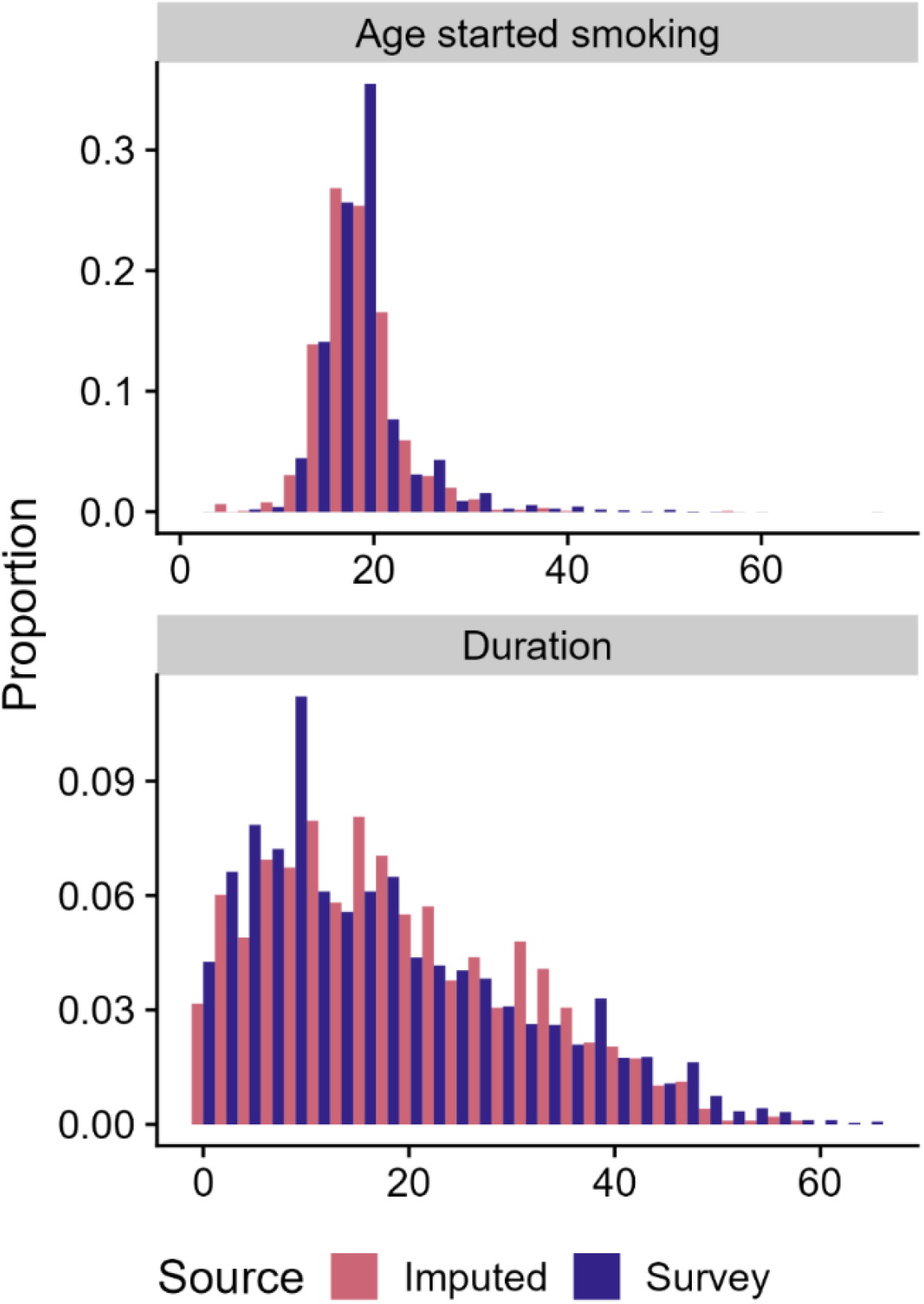
Comparison between imputed and observed smoking initiation and duration in the 2019 National Drug Strategy Household Survey.

